# Problem-based Learning Curriculum Disconnect on Diversity, Equity, and Inclusion

**DOI:** 10.1101/2024.02.02.24302186

**Authors:** Mario Brondani, Grace Barlow, Shuwen Liu, Pavneet Kalsi, Annika Koonar, Jialin (Lydia) Chen, Peter Murphy, Jonathan Broadbent, Bruna Brondani

## Abstract

**Background:** Diversity, equity, and inclusion (DEI) mission statements continue to be adopted by academic institutions in general, and by dental schools around the globe in particular. But DEI content seems to be under-developed in dental education.

**Objectives:** The objectives of this study were two-fold: to extract information from all the PBL cases at University of British Columbia’s Faculty of Dentistry curriculum in terms of the diversity, equity, and inclusion of patient and provider characteristics, context, and treatment outcomes; and; to compare these findings with the composition of the British Columbia census population, dental practice contextual factors, and the evidence on treatment outcomes within patient care.

**Methods:** Information from all the 58 PBL cases was extracted focusing on patient and provider characteristics (e.g., age, gender, ethnicity), context (e.g., type of insurance), and treatment outcomes (e.g., successful/unsuccessful). This information was compared with the available literature.

**Results:** From all the 58 PBL cases, 0.4% included non-straight patients, while at least 4% of BC residents self-identify as non-straight; there were no cases involving First Nations patients although they make up 6% of the British Columbia population. Less than 10% of the cases involved older adults who make up almost 20% of the population. Only Treatments involving patients without a disability were 5.74 times more likely to be successful compared to those involving patients with a disability (*p*<0.05).

**Conclusions:** The characteristics of the patients, practice context, and treatment outcomes portrayed in the existing PBL cases seem to differ from what is known about the composition of the British Columbia population, treatment outcome success, and practice context; a curriculum disconnect seems to exist. The PBL cases should be revised to better represent the population within which most students will practice.

## INTRODUCTION

Diversity, equity, and inclusion (DEI) mission statements continue to be adopted by academic institutions in general, and by dental schools around the globe in particular [1,2,3,4]. The term “diversity” is usually used to refer to differences among individuals, including but not limited to race, ethnicity, disability, nationality, socioeconomic stratum, gender identity, and sexual orientation [5]. The term “equity” refers to the goal of fair and unbiased treatment and representation of all individuals of a group or community regardless of their diversity. The term “inclusion” refers to the extent to which any individual or group are or feel respected, supported, and engaged in a given environment [6]. Accordingly, DEI statements within dental schools focus solely on people—students, staff, faculty members, and patients—so they feel safe, welcomed, valued, represented, and heard [7]. In turn, there are a number of initiatives aimed at increasing representation of students, patients, staff, and faculty from various backgrounds in dental education [8,9], and to encourage DEI discussions aimed at improving cultural sensitivity within undergraduate training [10].

It is indeed imperative to ensure that diverse individuals are equally represented within faculty, student, staff and student bodies, and it is also important to show this diversity in conjunction with equity and inclusion within the didactic content with which students are taught, from pedagogical material to teaching cases and essays [11]. DEI principles have been integrated into a number of medical curricula around the world [12,13], but this seems to be under-developed in dental education generally [14]. Dental education employs an array of teaching methods, including in-person lectures, recorded and lectures, and large and small group discussions of clinical problems and cases portraying dental-related scenarios, written essays and assignments, and others [15,16,17,18].

In the Faculty of Dentistry (FoD)at the University of British Columbia (UBC), the four-year undergraduate dental curricula utilise various teaching methods, one being hybrid problem-based leaning (PBL) curriculum [19]. In the hybrid PBL, the weekly clinical problem portrayed in the scenario is discussed in a small-group learning activity with around eight students monitored by a tutor, while part of the didactic content related to the problem at-hand is delivered via lectures, allowing students to move beyond memorising the information to then restructure the facts into knowledge by combining their self-learning in small groups with the lecture material [20]. Two courses that employ the largest number of PBL in the curriculum at the UBC’s FoD are Fundamental Medical Sciences (FMS) I and II in Years 1 and 2 of the undergraduate dental program, respectively. Although these PBL cases were introduced in 1997, when undergraduate dental students studied the fundamentals of medical sciences jointly with undergraduate medical students, a 2010 study highlighted the need to redesign and redirect the objectives of this basic curriculum to make it more relevant to dental professionals [21]. Such 2010 study led to a curriculum review which culminated with the Faculty of Dentistry teaching undergraduate dental students separately from Faculty of Medicine students starting in the 2015-16 academic year. The PBL cases have since been revised in terms of their dental and oral health relevance and content, but it is unlikely that these revisions addressed issues of diversity, equity, and inclusion in regard to patient and provider characteristics, context, and treatment outcomes. Diversity and inclusive teaching is essential for preparing civically engaged health care providers and for creating a society that recognizes the contributions of all people [22], and should be reflected on the teaching material we use so that we acknowledge a range of differences in patients and cases, embrace such differences, and allow these differences to transform the way we think, teach, learn and act.

The objectives of this study were two-fold: 1) to extract information from all the PBL cases at UBC’s FoD curriculum in terms of the diversity, equity, and inclusion of patient and provider characteristics, context, and treatment outcomes; and 2) to compare these findings with the composition of the British Columbia census population, dental practice contextual factors, and the evidence on treatment outcomes within patient care. Our research question was: “To what extent do the University of British Columbia’s Faculty of Dentistry PBL cases represent the composition of the BC population in terms of patient characteristics; the context of the profession in terms of provider, dental insurance, and dental practice characteristics; and the various treatment outcomes based on the existing literature?”.

## METHODS

After gathering all 58 PBL cases from Fundamental Medical Sciences (FMS) I and II, the two case-based courses in Years 1 and 2 of the undergraduate dental curriculum at UBC’s Faculty of Dentistry, an extraction table was developed (Table 1). The information was extracted from each PBL case by two trained researchers, and was based on:

**Table 1.** Patient, treatment outcome, and dental practice characteristics portrayed in the cases compared to the general population composition, evidence of treatment outcome success, and types of dental practices in British Columbia, Canada.

| *Patient, treatment outcome and dental practice characteristics* | *In the PBL cases<sup>^</sup>* | *In the XXX population, in the evidence around treatment outcomes, and in the types of dental practices* |
| --- | --- | --- |
| Sexual orientation of patients:<br>*non-straight* | ➤ **1.7%** of cases (1 case) portrayed non-straight patients | ➤ **4%** of XXX are members of the 2SLGBTQIA+** community |
| Age of patients:<br>*older than 65 years of age* | ➤ **8.6%** of cases (5 cases) portrayed patients older than 65 | ➤ **19%** of XXX are older than 65<sup>25</sup> |
| Patients' body portrayal:<br>*Living with a disability* | ➤ **13.7%** of cases (8 cases) included patients younger than 64 years of age with a disability | ➤ **20.5%** of XXX between the ages of 15 and 64 live with a disability<sup><</sup> |
| *Overweight* | ➤ **12%** of cases (7 cases) showed people who were overweight | ➤ **23%** of XXX are overweight |
| Race/ethnicity of patients:<br>*First Nation Peoples* | ➤ **0%** of cases involved First Nations Peoples | ➤ **6%** of XXX are of Indigenous heritage |
| *Visible minorities\** | ➤ **6.8%** of case (4 cases) patients were from a visible minority group | ➤ **30%** of XXX are from a visible minority group<sup>22,23</sup> |
| Language spoken by the patient and the provider:<br>*English* | ➤ **100%** of the patients spoke/understood English | ➤ **3.3%** of XXX do not speak English nor French |
|  | ➤ But **3.4%** of cases (2 cases) had patients that seemed to have English as a second language | ➤ **28%** of XXX do not have either English or French as their official language<sup><</sup> |
| Treatment success:<br>*Successful* | ➤ **91.3%** of cases (53 cases) had treatments that seemed to be successful | ➤ The success rate can be as low as **50%** for fillings<sup>50</sup> and **60%** for root canal treatment over time<sup>51</sup> |
| Affordability:<br>*Able to pay for the treatment provided* | ➤ **13.7%** of the cases (8 cases) mentioned inability to afford treatment (surrogate for not having insurance) | ➤ **16%** of XXX cannot afford dental care, **25%** in XXX<sup>24</sup> |
| Dental practice characteristic:<br>*University dental clinic* | ➤ **10.3%** of cases (6 cases) were at the XXX dental clinic | ➤ **4%** on the dental practices are corporations, **37%** are sole practitioners, and **32%** are shared practices<sup>46,48</sup> |
| *Private practice* | ➤ **63.7%** of cases (37 cases) were in private (sole) practice |  |
| Providers | ➤ **40%** of cases (23 cases) showed the students as the providers either as associates (15 cases) or owners (7 cases) |  |

^ Includes all the 58 Problem-Based Learning cases.
\* In XXX, the Employment Equity Act defines visible minorities as persons, other than Aboriginal peoples, who are non-Caucasian in race or non-white in colour, and consists mainly of the following groups: South Asian, Chinese, Black, Filipino, Latin American, Arab, Southeast Asian, West Asian, Korean and Japanese.<sup>23</sup>
\*\* The acronym considers sexual orientation, gender identity and gender expression. 2S refers to Two-Spirit People in the First Nation and Indigenous culture; L refers to Lesbian; G refers to Gay; B refers to Bisexual; T refers to Transgender; Q refers to Queer; I refers to Intersex, and + is inclusive of people who identify as part of sexual and gender diverse communities, who use additional terminologies.
< https://www2.gov.bc.ca/gov/content/home/accessable-government/toolkit/audience-diversity

- Patient characteristics (age, gender, sexual orientation, ethnicity, language spoken, and body type);
- Provider characteristics (age, gender, sexual orientation, and ethnicity);
- Context (type of insurance if any, type of dental practice of the provider including private and hospitals);
- Treatment outcome (successful, unsuccessful).

The extraction table focused on diversity (e.g., the variety of characteristics among cases pertaining to patients, providers, context, and treatment outcomes); equity (the fair distribution of these differences across the PBL cases); and inclusion (e.g., the extent to which these differences were represented across cases). These characteristics were compared with the demographic composition of the British Columbia census population (note that British Columbia is a province with high ethnic diversity). We also compared the content of the PBL cases with the dental practices’ ownership characteristics (sole/private, group practice, corporation, etc.), patients’ ability to understand and afford care, and with the evidence of success of various treatment outcomes based on existing literature. Characteristics relating to the composition of the province’s general population were obtained through official government data [23,24] and our previous work [25]. All data collected were exported into IBM® SPSS (Version 27) software for statistical analysis. Descriptive statistics included summaries of the data in terms of means and frequencies. The Pearson’s chi-square test (odds ratio) was utilised at *p* < 0.05.

## RESULTS

Upon reviewing the curriculum, a total of 58 PBL cases were identified for inclusion and analysis in this study. These 58 cases make-up all the PBL material available and included 30 cases for FMS I and 28 for FMS II. Table 1 shows the percentages of patients, treatment outcomes, and dental practice’s characteristics portrayed in the PBL cases and contrasts this with the British Columbia population composition, evidence of treatment outcome success rates, and the types of dental practices in the province.

Figure 1 offers a comparison between those with and without insurance in the PBL cases and in the general population. Treatments involving patients without a disability were 5.74 (95% CI [1.28, 32.7], *p*=0.04) times more likely to be successful than those involving patient with a disability. Cases involving patients unable to afford treatment were 2.81 (95% CI [0.42,19.21], *p*=0,5) times more likely to be referred to another dentist/clinic than those able to pay/with insurance.

**Figure 1.**
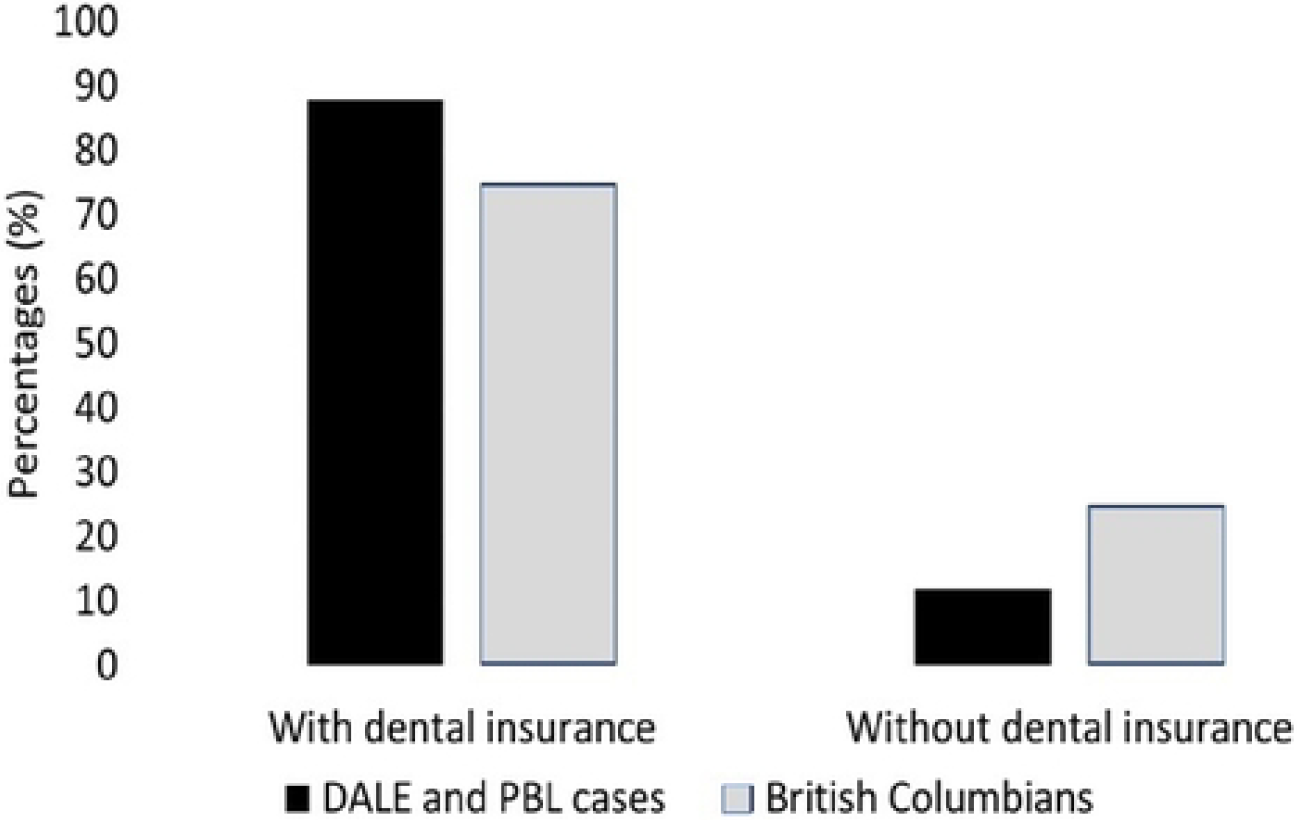
Dental Insurance for British Columbia and the PBL cases.

## DISCUSSION

This study was the first to situate all the cases in the form of PBL from the two largest problem-based courses at UBC’s undergraduate dental program in terms of their diversity, equity, and inclusion. In particular, we answered to our research question by exploring the extent to which patients’ and providers’ characteristics, context, and treatment outcomes were represented within the cases when compared to the population’s characteristics, dental practice contextual factors and the evidence of successful treatment outcomes within patient care. We found that more work is needed in this regard. For example, the PBL cases seemed to accurately reflect the prevalence of disability among British Columbians according to its Government [26], although obesity was also referred to as a disability throughout the cases without any attempt to discuss its interplay with disease that can lead to increased weight-related stigma as found by Hilbert and colleagues [27]. The PBL cases also seemed to realistically emphasise that around 16% of Canadians are unable to afford the recommended dental care treatment although that percentage is much higher - 25% - among British Columbians who would not likely be able to afford the treatment offered and delivered within the cases [25]. Also, dentists’ diversity could have been implied when 23% of providers portrayed in the cases were the students themselves (*“You are the attending dentist when the patient arrives*…*”*), representing whoever they are in terms of gender identity, race, ethnicity, and other socio-demographic characteristics. But, in general, despite efforts to include diversity, equity, and inclusion mission statements on dental school websites worldwide, PBL teaching material at the UBC’s Faculty of Dentistry seems to fall short on these efforts, according to our findings, showing a disconnect between diversity and inclusive teaching in the classroom and diversity and inclusive teaching in the didactic material employed in the curriculum.

It is well known that problem-based learning methodology can enable students to develop superior professional skills and effective learning strategies compared with those instructed using traditional approaches such as lectures [28,29]. However, the discrepancy between most of the overall characteristics of the cases discussed within this PBL curriculum and the context in which graduates will be practicing in our study seems to attest to the lack of authenticity in utilizing a more realistic health educational environment for the students regardless of the teaching pedagogy employed, as reported by Lee and colleagues when discussing health-related training [30]. Although there is evidence that fresh graduates may feel relatively unprepared [31] to perform certain clinical procedures after they graduate [32,33,34,35,36], more realistic cases are still needed to better represent patients, regardless of clinical procedures to be performed.

The technical skills required to perform a given treatment do not vary based on patients’ skin colour, sexual orientation, or body type, but interpersonal skills and understanding may vary based on patients’ socio-economical and personal differences. Therefore, it is paramount to have these procedures contextualized within these characteristics and patient’s diverse backgrounds and contexts. For example, by not fully representing the array of patient ethnicities, the PBL cases might inadvertently overlook the importance of cultural practices in shaping peoples’ oral health behaviours [37,38], particularly for Indigenous Canadians [39,40] who were not represented at all in the cases. Such overlook may hinder the understanding of Indigenous cultural competency that is needed to recognize, comprehend and appreciate the values, traditions and belief systems of Indigenous peoples that may be markedly different from one’s own. And by not representing languages other than English or French that are spoken in the Province by many patients, the PBL cases might have downplayed the relevance of establishing patients’ rapport and their abilities to fully understanding treatment options so that informed consent is indeed fully obtained [41,42].

As affordability for some remains a driver to receiving recommended dental treatment and having dental insurance is a major facilitator in doing so [43], the PBL cases should more realistically portray patients who are not able to afford dental care in the province, and who do not have insurance (Figure 1). Around 32% of Canadians and more than 30% of British Columbians have no dental insurance [25], and although insurance was mentioned in some of the cases, it was not identified as a key factor for patients obtaining the care they needed, downplaying the impact of affordability to more than 25% of British Columbians. But at the same time, the PBL cases should now better reflect the potential impact of the newly introduced Canadian Dental Plan [44] that is designed to help ease financial barriers to accessing oral health care for uninsured Canadian residents bellow an annual family income of less than CAD$90,000 (US$: 66,500 on February 1, 2024).

With most patients in the PBL cases being portrayed as relatively younger (less than 9% were older than 65 years), students might not fully understand the impact of aging in the provision of oral health care when almost 20% of the population is already older the 65 and some might present with systemic health issues, comorbidities and limitations [45]. Although the students do learn about aging in subsequent years of their training [31,46], it is important to better represent our ever growing older adult population in all aspects of our teaching and to avoid biased views of aging that might lead to ageism and prejudices against older adults as we discussed previously [47].

Over the years the landscape of practicing dentistry has changed from the dominant – and traditional model of sole providers to more group practices and even large corporations [48,49,50]. In turn, the PBL cases could reflect such changes in landscape to better mediate future graduates’ expectations about their working environment. The cases should also offer a more realistic expectation about successful treatment outcomes below the 90% success rate currently presented, as failures and re-dos do occur over time, are part of the practice of dentistry [51,52,53,54] and should be discussed accordingly for a more realistic practicing scenario.

Lastly, the unfortunate associations we found showing treatments involving patients without disability being more likely to be successful than those involving patients with a disability, and cases involving patients unable to afford treatment being more likely to be referred to another dentist, might send the wrong ethical and professional message. Patients with a disability and those unable to afford care already face adversities when accessing oral health care [55,56] and the association portrayed herein may not only perpetuate these experiences, but also impede efforts to teach social responsibility [17] and ethics in health care [16].

Despite its findings, this study has limitations. The focus on one dental school prevents generalisations to other curricula. Given that PBL cases are not the only source of learning, students might have been exposed to diversity, equity, and inclusion elsewhere in their dental training which was not considered in this study. The PBL cases are facilitated by a tutor—usually a volunteer dentist—and we do not know if these tutors might have incorporated their diverse views when discussing the cases with the students. Whether or not patients portrayed in teaching cases are fictitious or real, they must reflect the composition and characteristics of the population graduates will serve. As the PBL cases should be revised to better reflect these characteristics, future studies should assess the extent to which these changes impact students’ understanding and appreciation for diversity, equity, and inclusion in oral health care. It is also important to empower students to engage in these types of curriculum revisions aimed at improving their own education through collaborative input, as suggested by French and colleagues [57], while respecting their busy schedules to avoid burn out.

## CONCLUSIONS

The characteristics of the patients, practice context, and treatment outcomes portrayed in the existing PBL cases seem to differ from what is known about the composition of the British Columbia population, treatment outcome success, and practice context; a curriculum disconnect seems to exist. Although dentists’ diversity could be implied when 23% of the students were portrayed as the providers in the cases representing their own characteristics, the existing PBL curriculum should be revised to better portray the mosaic of our society and treatment outcomes. Students should have more opportunities to actively think about the barriers to care that other groups and populations face based on their inherent characteristics, and to refrain from stereotyping certain populations in practice. The PBL cases should be revised to better represent the population within which most students will practice.

## Data Availability

Full data cannot be shared publicly because they are part of a series of PBL didactic cases from the UBC's FoD dental curriculum, and belong to the creators - instructor. Partial data are available from the first author (MB) as one of the creators of the PBL cases.

## ABBREVIATIONS

BC: British Columbia
CI: Confidence interval
DEI: Diversity, Equity and Inclusion
FMS: Fundamentals of Medical Science
FoO: Faculty of Dentistry
PBL: Problem-based Learning
UBC: University of British Columbia

## DECLARATIONS

### Ethical approval and consent to participate

Please note that this study does not involve the recruitment of human participants, so there is no ethics to be approved.

### Consent for publication

Not applicable

### Availability of data and materials

Full data cannot be shared publicly because they are part of a series of PBL didactic cases from the UBC’s FoD dental curriculum, and belong to the creators-instructor. Partial data are available from the first author (MB) as one of the creators of the PBL cases.

### Authors’ contributions

MB contributed to the study conception, design, data acquisition and interpretation, as well as drafting and critically revising the manuscript. BB and LC contributed to the mapping of the cases and organized the references. GB, SC, PK, and AK contributed to data interpretation and organizing the table and figure. PM and JB contributed to writing and revision of the manuscript.

## ACKNOWLEDGMENTS

The authors acknowledge the report provided by the Students for Collective Change in 2021 that outlined the possible diversity, equity, and inclusion shortfalls within the existing PBL cases. This study received financial support from the 2023 University of British Columbia Students as Partners initiative. An abbreviated version of this study was presented during the 101st International Association for Dental Research Annual Conference in Bogota, Colombia, on June 23, 2023.

